# Preliminary Reliability and Validity of SynapTrack, a Smartphone-Based Digital Biomarker Platform for Remote Assessment of Cervical Spondylotic Myelopathy

**DOI:** 10.64898/2026.05.29.26354454

**Authors:** Salim Yakdan, Pranay Singh, Faraz Arkam, Erdong Chen, Abigail Lewis, Benjamin Steel, Isaac Becker, Wenwen Guo, Haris Naveed, Carl Wang, Deyuan Yang, Zhuoran Wang, Wilson Z. Ray, Jason Hassenstab, Michael P. Steinmetz, Zoher Ghogawala, Caitlin Kelleher, Jacob K Greenberg

## Abstract

**Background and Objectives:** Cervical spondylotic myelopathy (CSM) is a leading cause of neurological disability in older adults. However, validated, scalable tools to quantify disease severity and changes over time are lacking. Recent advances in smartphone technology have opened new avenues for longitudinal, objective, and remote monitoring of neurological conditions. We performed a preliminary evaluation of the reliability and validity of SynapTrack, a smartphone-based digital platform for objective remote CSM assessments.

**Methods:** In this single-center prospective cohort study, 251 participants (151 with CSM, 100 healthy controls) completed in-person SynapTrack assessments related to tapping, pinching, and vibratory detection, along with reference laboratory measures of dexterity (Box and Block Test, 9-Hole Peg Test) and vibratory sensation (tuning fork). A subset completed repeated home-based testing to assess test-retest reliability. We evaluated convergent validity, construct validity against the modified Japanese Orthopedic Association (mJOA) score, known-groups validity, and test-retest reliability (intraclass correlation coefficient, ICC).

**Results:** Smartphone-derived metrics demonstrated good-to-excellent test-retest reliability, with the strongest stability for vibratory detection threshold (ICC = 0.92), overall and non-dominant tapping speed (ICC = 0.90 each), and pinching successful targets (ICC = 0.90). Convergent validity was supported by moderate-to-strong correlations between digital metrics and reference laboratory dexterity tests (ρ up to 0.60 for tapping speed; up to −0.65 for the vibratory threshold). Construct validity against the mJOA was strongest for the vibratory threshold (ρ = −0.53 to −0.54) and Level 2 non-dominant pinching errors (ρ = −0.45). Selected metrics distinguished CSM patients from controls with good discrimination, including non-dominant tapping speed (AUROC = 0.76, 95% CI 0.68–0.85), Level 2 dominant pinching successful targets (AUROC = 0.78, 95% CI 0.62–0.94), and the non-dominant vibratory threshold (AUROC = 0.77, 95% CI 0.64–0.90).

**Conclusions and Relevance:** A smartphone-based battery of upper-extremity sensorimotor tasks demonstrated preliminary reliability and validity in CSM. Furthermore, to our knowledge, the novel vibratory detection task represents the first smartphone-based sensory assessment used for CSM. Collectively, these findings position SynapTrack as a scalable platform for objective, remote neurological monitoring of CSM.

## INTRODUCTION

Cervical spondylotic myelopathy (CSM) is a major cause of neurological disability, particularly among older adults. It results from chronic cervical spinal cord compression due to age-related degenerative changes and represents the most common cause of spinal cord dysfunction in this population.^1^ Although the precise prevalence is difficult to define, the best available evidence suggests that CSM affects approximately 2% of adults.^2^ Even more common is its antecedent prodromal phase of asymptomatic spinal cord compression (SCC) identified on cervical imaging.^2^ For example, radiographic SCC is present in approximately one-third of asymptomatic adults older than 60 years, and up to 25% of these individuals may eventually develop symptomatic myelopathy.^2–6^ As the population ages, the prevalence and impact of both SCC and symptomatic CSM are therefore expected to grow.

CSM can cause a range of disabling neurological impairments, most commonly impaired dexterity and gait dysfunction, but also weakness, balance impairment, and bladder dysfunction.^4,7^ Beyond its neurological manifestations, CSM is associated with substantial declines in physical function and quality of life.^8^ Surgery remains the only treatment with high-level evidence for halting disease progression and improving neurological function.^9^ However, because surgery carries meaningful risks despite generally more favorable outcomes with earlier intervention,^10,11^ mild CSM and asymptomatic SCC are typically observed initially, with neurological progression serving as a key trigger for surgical intervention.^12,13^ In routine practice, this progression is typically monitored through patient report and intermittent clinician evaluations once or twice per year.^14^ The most widely used disease-specific instrument for monitoring CSM severity is the modified Japanese Orthopedic Association (mJOA) score, which is most commonly assessed based on patient report and does not directly quantify objective neurological function such as dexterity.^15–19^ The mJOA also has limited discrimination, only moderate reliability, and weak associations with broader measures of disability and quality of life.^14,20,21^ Improved objective tools capable of detecting early or subclinical neurological decline could support more personalized surgical decision-making by better balancing operative risks against the potential for irreversible progression. In fact, improving such monitoring tools has been formally prioritized as a key research goal for CSM by the AO Spine RECODE-DCM initiative.^22^

Advances in mobile health (mHealth) and smartphone technology have created new opportunities to address this gap. Smartphone-based digital biomarkers are objective, sensor-derived measures of physiological or behavioral function that can be captured repeatedly and remotely, outside the clinic and with minimal burden.^23^ Examples include quantifying manual dexterity from touchscreen-based tapping tasks or gait from built-in accelerometers and gyroscopes.^24,25^ Such assessments have already shown promise in neurological conditions such as multiple sclerosis and Parkinson’s disease, highlighting a compelling opportunity to develop similar tools for CSM.^24–27^

To address this unmet need, we developed SynapTrack, a smartphone-based digital platform designed to provide objective assessments of neurological function in patients with CSM through structured testing of gait, dexterity, and vibratory sensation.^28^ In this study, we aimed to perform a preliminary evaluation of the reliability and validity of smartphone-derived digital metrics collected through SynapTrack.

## METHODS

### Study Population

This single-center prospective cohort study was conducted at a tertiary academic medical center. Eligible participants included English-speaking adults aged 18 years or older with a confirmed diagnosis of CSM established by a fellowship-trained spine surgeon based on clinical presentation and supporting cervical imaging demonstrating spinal cord compression. Healthy controls without a history of cervical myelopathy or any other known condition affecting their upper extremity function were also recruited for comparison. Exclusion criteria included major comorbid neurological disorders that could confound neurological assessment (e.g., Parkinson’s disease, stroke, traumatic brain injury), severe cognitive impairment limiting ability to follow study instructions, significant musculoskeletal or physical limitations preventing completion of study testing, substantial visual, auditory, or motor impairments unrelated to CSM that could interfere with app use, and non-degenerative causes of myelopathy (e.g., tumor, infection, inflammatory disease). The study was approved by the institutional review board (IRB #202401026), and written informed consent was obtained from all participants.

### SynapTrack

SynapTrack is a custom smartphone application to test neurological abilities available on iOS and Android,^28^ though most participant testing occurred using the iOS platform. SynapTrack includes tests of dexterity through rapid tapping and pinching tasks, sensory/vibratory detection, and gait quality. In this manuscript, we focus on the upper extremity tasks. Notably, SynapTrack was developed iteratively over time, including working through earlier versions of drawing/pinching and vibratory tasks to arrive at the current refined versions. Therefore, the number of participants who completed each task varied according to the timing of its development. Representative images from SynapTrack are shown in Figure 1A-D.

**Figure 1.**
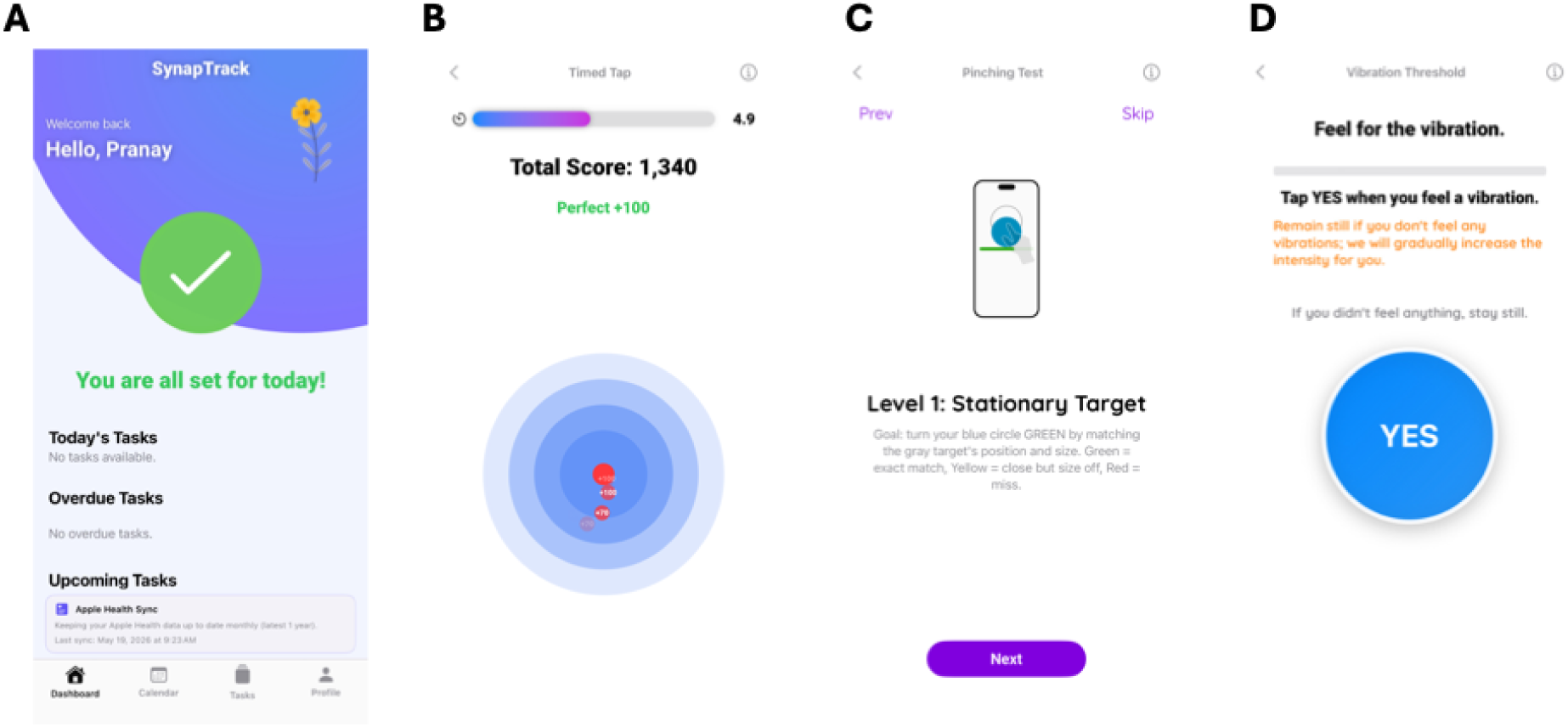
Representative screens from the SynapTrack iOS application. (A) Home dashboard displaying the participant’s daily task status and upcoming assessment schedule. (B) Timed tapping task, in which participants rapidly tap a target while accuracy- and speed-based scores accumulate. (C) Pinching task, in which participants pinch and drag a shape to match the size and position of a target (Level 1, stationary target shown). (D) Vibration threshold task using an adaptive staircase procedure, in which stimulus intensity is incrementally increased and participants tap “YES” upon perceiving a vibration.

### Data Collection

All participants underwent a standardized in-person baseline assessment; some completed more detailed, conventional laboratory-based neurological and functional testing, while others underwent briefer testing within a clinic setting. Reference clinical assessments were selected to capture domains relevant to CSM, including upper extremity dexterity and sensory function. Upper extremity dexterity was evaluated using established performance-based measures, including the Box and Block Test and the 9-Hole Peg Test. Sensory function was assessed using standardized vibratory testing with a tuning fork. Disease severity and patient-reported functional status were also assessed using validated clinical instruments, including the mJOA score and relevant patient-reported outcome measures.

Following laboratory testing, participants completed SynapTrack assessments on a smartphone under supervised conditions. Participants completed an initial supervised practice session, followed by tasks that were later included in core performance assessments. A subset of participants subsequently completed home-based testing over multiple days to evaluate test-retest reliability of remote digital assessments. Over the course of testing, tasks were modified and new tasks were developed; therefore the number of participants completing each task varied accordingly.

### SynapTrack upper-extremity and sensory tasks

Each task was administered separately for the dominant and non-dominant hand.

#### Tapping

Participants used their index finger to tap a circular on-screen target as rapidly and accurately as possible during a fixed 10-second interval. Taps at the center of the target received full credit, while taps falling progressively farther from the center received reduced credit, rewarding both speed and spatial precision. Two principal metrics were derived: tapping speed, the rate of registered taps over the trial, and tapping positional error, the mean spatial deviation of taps from the target center (with higher values reflecting reduced spatial precision).

#### Pinching

Using the thumb and index finger, participants dragged a blue circle to a target location and pinched to resize it so that its diameter matched a target ring. When the resized circle matched the target within a narrow tolerance it was displayed green (a successful trial); when it was slightly larger or smaller it was displayed yellow. If outside of the target, it was displayed red. The task comprised two levels of 10 trials each. In Level 1 the target remained in a fixed position across trials; in Level 2 the target appeared in a new position on each trial. Derived metrics included the number of successful targets (those matched within the target tolerance, displayed green), the number of error targets (those falling outside the target, displayed red), and total completion time.

#### Vibratory detection threshold

Participants held the smartphone in the hand being tested while the device delivered a vibratory stimulus that increased from zero in fixed increments. Participants pressed a large “Yes” button as soon as they perceived vibration. Stimulus intensity followed an adaptive staircase with equal step sizes that reversed direction at each perceptual transition (detection to non-detection, or vice versa).^29^ The vibratory detection threshold was computed as the mean of the staircase reversal points.

Higher thresholds indicate poorer vibratory sensitivity.

### Statistical Analysis

#### Descriptive Statistics

Participant characteristics were summarized using descriptive statistics. Continuous variables were reported as mean ± standard deviation (SD) or median with interquartile range (IQR), as appropriate based on distributional characteristics, while categorical variables were summarized as frequencies and percentages. Group comparisons between CSM patients and healthy controls were performed using independent-samples t-tests or Wilcoxon rank-sum tests for continuous variables and chi-square or Fisher’s exact tests for categorical variables, as appropriate.

### Convergent Validity

Convergent validity was assessed by examining associations between SynapTrack-derived digital metrics and established laboratory-based measures assessing similar neurological domains. Specifically, upper extremity digital biomarkers were compared with conventional dexterity and sensory assessments including the Box and Block Test, 9-Hole Peg Test, and standardized clinical vibratory testing. Pearson or Spearman correlation coefficients were calculated depending on data normality. Correlation strength was interpreted using standard conventions, with coefficients of approximately 0.30, 0.50, and 0.70 representing weak, moderate, and strong associations, respectively.^30^

### Construct Validity

Construct validity was assessed by examining associations between SynapTrack digital biomarkers and disease severity measures in the CSM cohort. Correlations between digital biomarkers and the mJOA score were evaluated using Pearson or Spearman correlation coefficients as appropriate. These analyses assessed whether digital metrics reflected patient-reported neurological impairment.

### Known-Groups Validity and Discriminative Performance

Known-groups validity was assessed by comparing SynapTrack digital metric performance between CSM patients and healthy controls. Discriminative performance of individual digital biomarkers was assessed using receiver operating characteristic (ROC) curve analysis, with calculation of the area under the ROC curve (AUROC) and corresponding 95% confidence intervals to quantify the ability of digital measures to distinguish CSM patients from healthy controls.^31^

### Test-Retest Reliability

Test-retest reliability of SynapTrack digital biomarkers was assessed in participants who completed repeated home-based testing. To reduce the influence of day-to-day within-subject variability, digital metric values were averaged across the first two days of home testing and compared with the corresponding averaged values from the final two available testing days within the 30-day observation period, or the final two preoperative testing days if surgery occurred earlier. Reliability was quantified using intraclass correlation coefficients (ICC[2,1]) from a two-way random-effects absolute-agreement model. ICC values were interpreted using conventional thresholds, with values <0.50 indicating poor reliability, 0.50–0.75 moderate reliability, 0.75–0.90 good reliability, and >0.90 excellent reliability.^32^ All statistical analyses were performed using R software (R Foundation for Statistical Computing). Statistical significance was defined as a two-sided p-value <0.05.

## RESULTS

### Participant Characteristics

A total of 251 participants were included in the study, comprising 151 patients with CSM and 100 controls. Because the SynapTrack task battery was developed and refined iteratively during the study, individual tasks were introduced at different time points, and most participants completed only a subset of the full assessment battery. Consequently, analytic sample sizes varied substantially across task domains, ranging from approximately 35 participants for pinching assessments to 50 participants for vibratory sensory testing and over 100 participants for tapping tasks.

For the overall enrolled cohort, the mean age was 60.2 ± 12.9 years, with a mean age of 62.4 ± 11.1 years in the CSM cohort and 56.9 ± 14.6 years in controls. Overall, 57% of participants were female, with a similar sex distribution between groups (55.6% in the CSM cohort vs 59.0% in controls). The cohort was predominantly White (83.3%), closely reflecting the demographic composition of our institution’s elective practice.^33^ A summary of participant demographic and clinical characteristics is provided in **Table 1**.

**Table 1.**
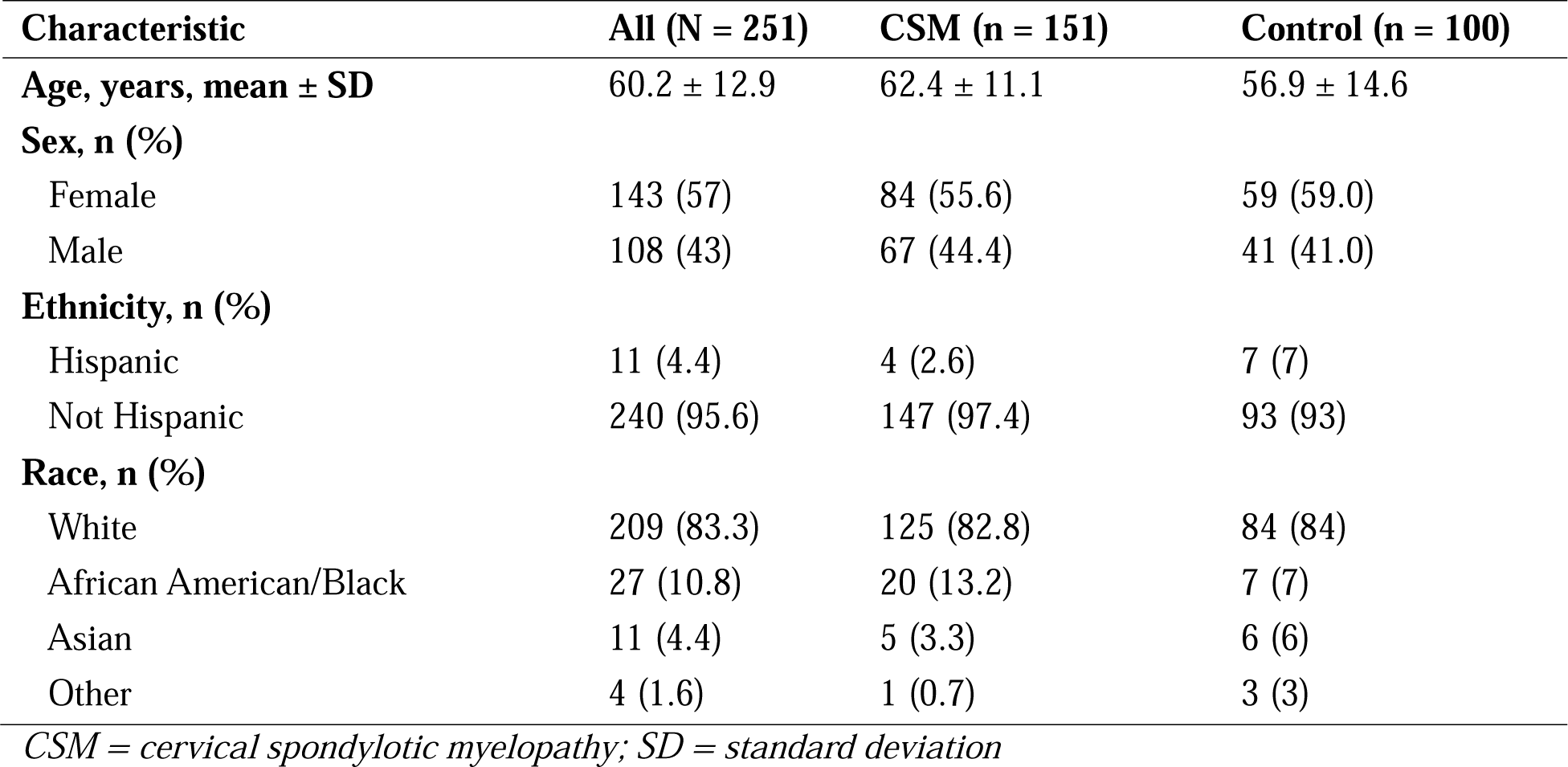
Participant demographic characteristics. Continuous variables are presented as mean ± standard deviation; categorical variables as n (%).

| Characteristic | All (N = 251) | CSM (n = 151) | Control (n = 100) |
| --- | --- | --- | --- |
| **Age, years, mean ± SD** | 60.2 ± 12.9 | 62.4 ± 11.1 | 56.9 ± 14.6 |
| **Sex, n (%)** |  |  |  |
| Female | 143 (57) | 84 (55.6) | 59 (59.0) |
| Male | 108 (43) | 67 (44.4) | 41 (41.0) |
| **Ethnicity, n (%)** |  |  |  |
| Hispanic | 11 (4.4) | 4 (2.6) | 7 (7) |
| Not Hispanic | 240 (95.6) | 147 (97.4) | 93 (93) |
| **Race, n (%)** |  |  |  |
| White | 209 (83.3) | 125 (82.8) | 84 (84) |
| African American/Black | 27 (10.8) | 20 (13.2) | 7 (7) |
| Asian | 11 (4.4) | 5 (3.3) | 6 (6) |
| Other | 4 (1.6) | 1 (0.7) | 3 (3) |

*CSM = cervical spondylotic myelopathy; SD = standard deviation*

### Convergent Validity

Tapping-derived digital biomarkers demonstrated strong convergent validity with established laboratory measures of upper-extremity dexterity (Figure 2). The strongest associations were observed for tapping speed metrics. Overall tapping speed demonstrated strong positive correlations with Box and Block Test performance (ρ = 0.58–0.60) and moderate inverse correlations with 9-Hole Peg Test completion time (ρ = –0.42 to –0.49). Similarly, non-dominant tapping speed showed strong correlations with Box and Block performance (ρ = 0.57 for both dominant and non-dominant Box and Block measures) and moderate inverse associations with 9-Hole Peg Test time (ρ = –0.47 to –0.53). Additional support was observed for dominant-hand tapping speed (ρ = 0.52–0.54 with Box and Block) and non-dominant tapping positional error, which correlated moderately with both Box and Block performance (ρ = –0.44 to –0.45) and 9-Hole Peg Test completion time (ρ = 0.50–0.51). Collectively, these findings support the ability of tapping-based digital biomarkers to capture clinically meaningful upper-extremity motor performance in patients with CSM.

**Figure 2.**
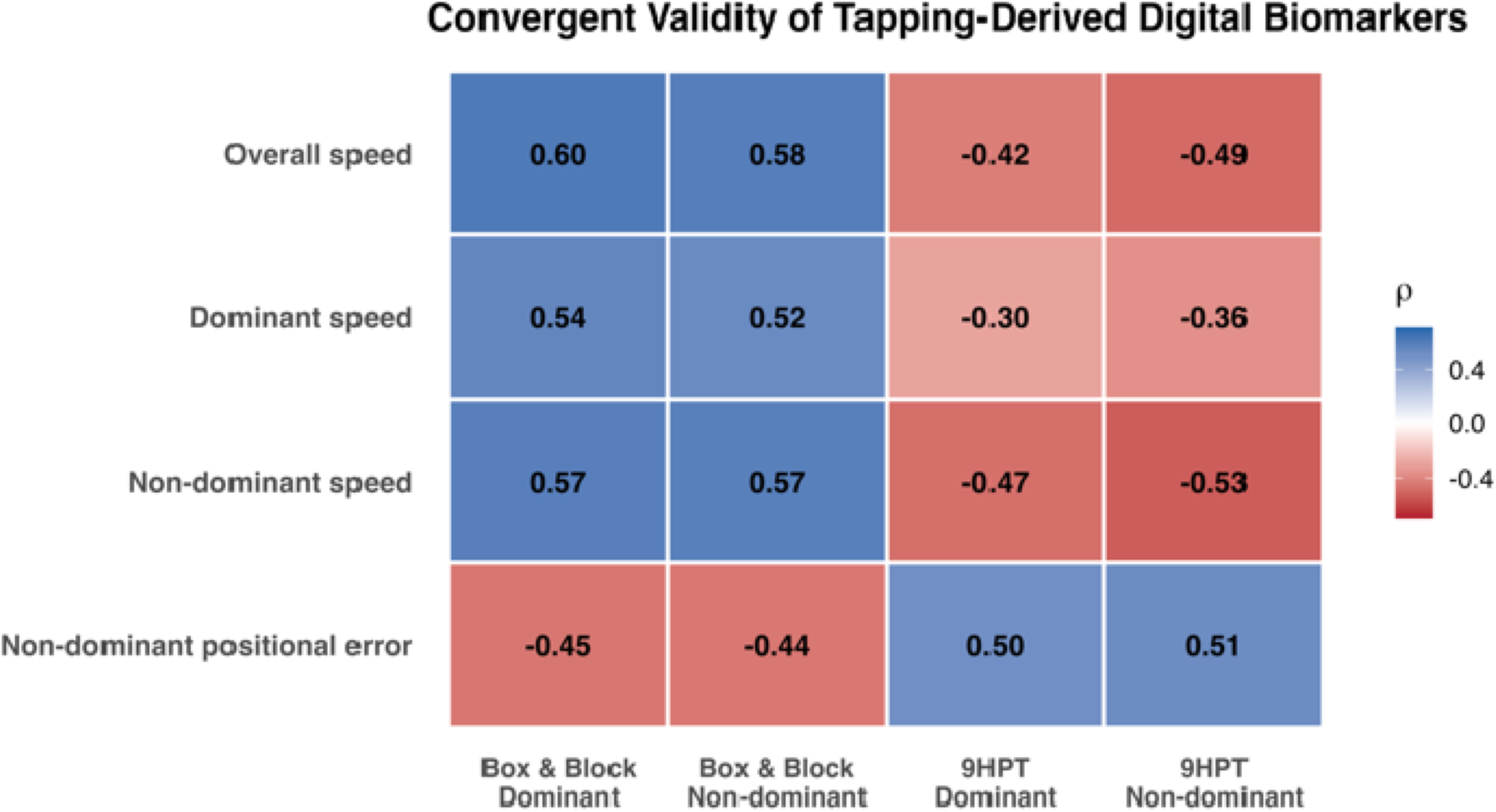
Convergent validity of tapping-derived digital biomarkers. Spearman correlation coefficients (ρ) between SynapTrack tapping metrics (overall, dominant, and non-dominant speed; non-dominant positional error) and reference laboratory measures of upper-extremity dexterity (Box and Block Test and 9-Hole Peg Test) for the dominant and non-dominant hands. Higher tapping speed and lower positional error are associated with better laboratory dexterity. 9HPT = 9-Hole Peg Test.

Pinching-derived digital biomarkers demonstrated consistent convergent validity with established laboratory measures of upper-extremity dexterity (Figure 3). The strongest and most consistent findings were observed for metrics reflecting task success and speed. The number of successful targets demonstrated moderate-to-strong associations with Box and Block performance and inverse associations with 9-Hole Peg Test completion time across both Level 1 and Level 2 conditions, with the strongest performance observed in the Level 2 dominant-hand task (ρ = 0.52–0.54 with Box and Block; ρ = −0.48 to −0.52 with 9-Hole Peg). Completion time also demonstrated consistent convergent validity, with slower task completion associated with poorer dexterity (ρ = −0.44 to −0.47 with Box and Block; ρ = 0.39–0.48 with 9-Hole Peg). Collectively, these findings suggest that SynapTrack pinching metrics capture clinically meaningful dexterity impairment across complementary domains of task success and speed.

**Figure 3.**
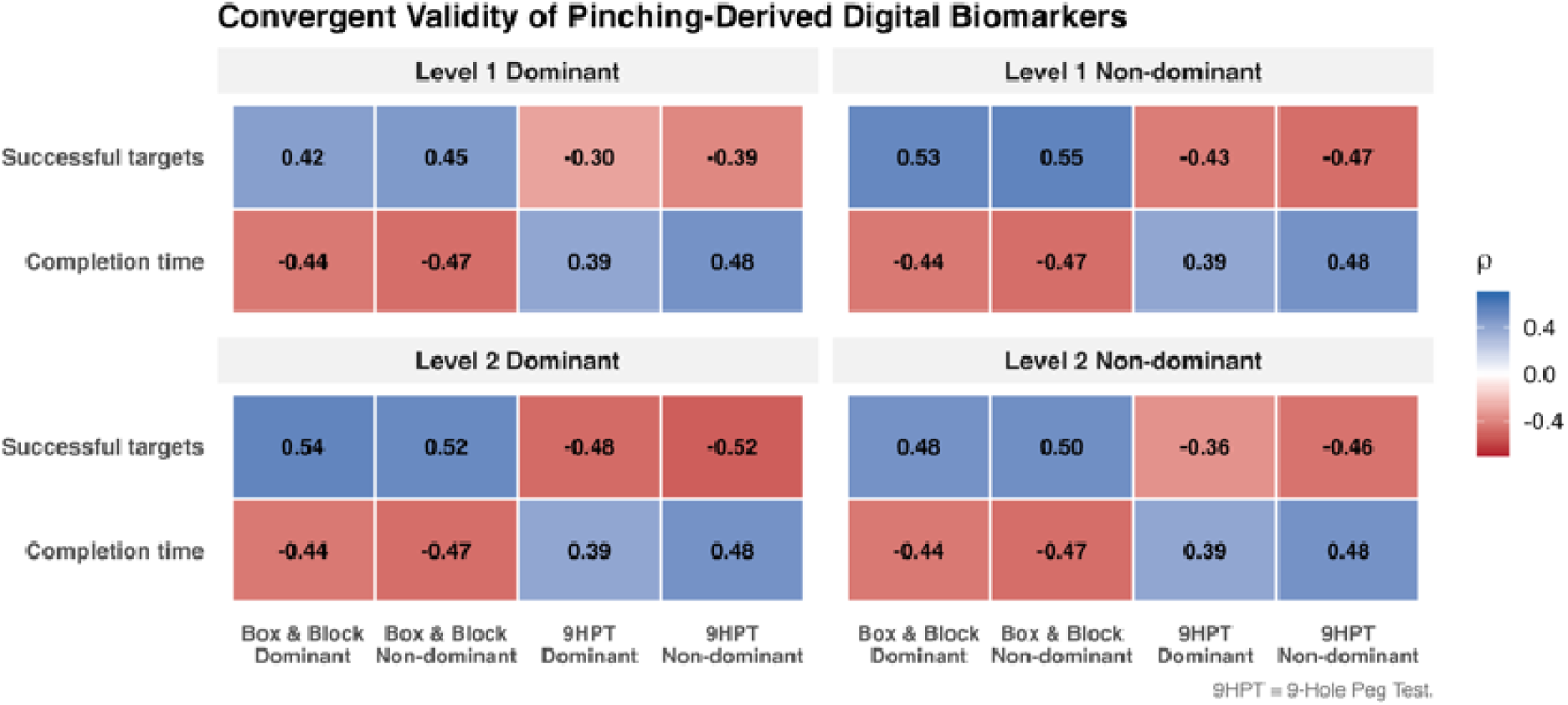
Convergent validity of pinching-derived digital biomarkers. Spearman correlation coefficients (ρ) between SynapTrack pinching metrics (number of successful targets and completion time) and reference laboratory dexterity measures (Box and Block Test and 9-Hole Peg Test), stratified by task level (Level 1: fixed target position; Level 2: shifting target position) and hand (dominant, non-dominant). Completion time reflects the total time required to complete the full pinching task battery across levels and therefore appears identically across conditions. 9HPT = 9-Hole Peg Test.

The smartphone-based vibratory detection threshold demonstrated consistent convergent validity with conventional neurological and dexterity assessments (Figure 4). Higher vibratory detection thresholds, reflecting worse sensory performance, were moderately associated with poorer upper-extremity dexterity on the Box and Block Test (ρ ranging from −0.49 to −0.65) and slower 9-Hole Peg Test performance (ρ ranging from 0.44 to 0.52). Importantly, the digital sensory metric also demonstrated moderate criterion validity relative to conventional vibratory testing with a tuning fork (dominant hand: ρ = −0.46, non-dominant hand: ρ = −0.44), supporting the ability of SynapTrack to capture clinically meaningful sensory impairment.

**Figure 4.**
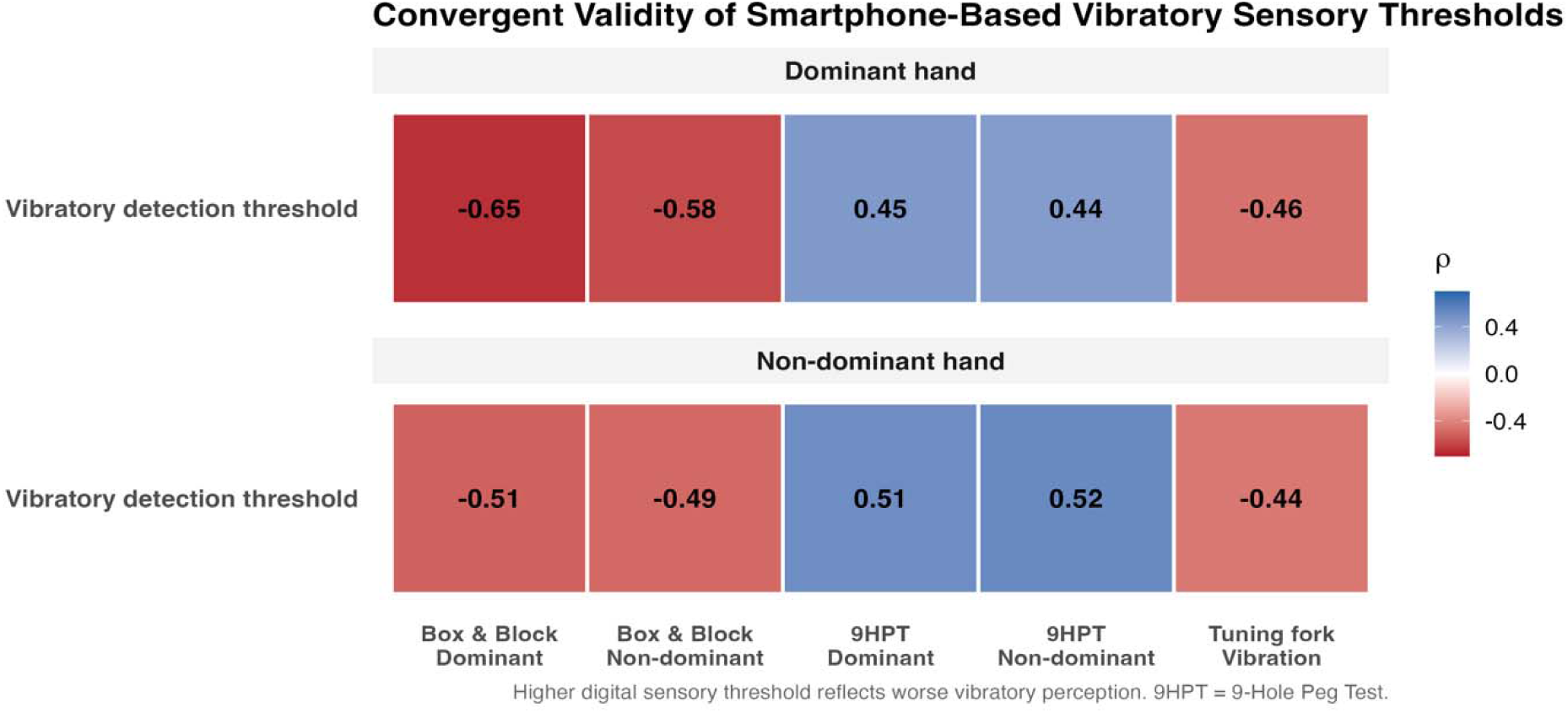
Convergent validity of the smartphone-based vibratory detection threshold. Spearman correlation coefficients (ρ) between SynapTrack vibratory detection thresholds (dominant and non-dominant hand) and reference laboratory measures of dexterity (Box and Block Test, 9-Hole Peg Test) and conventional clinical vibratory testing with a tuning fork. Higher digital sensory thresholds reflect poorer vibratory perception. 9HPT = 9-Hole Peg Test.

### Construct Validity

Tapping-derived digital biomarkers demonstrated modest but consistent association with patient-reported CSM disease severity (Figure 5). The strongest associations were observed for non-dominant tapping performance, particularly speed and positional error metrics. Non-dominant tapping positional error demonstrated a moderate inverse association with total mJOA score (ρ = −0.32), indicating that worse neurological impairment was associated with greater positional error (i.e., lower tapping precision). Non-dominant tapping speed also demonstrated modest associations with disease severity (total mJOA score, ρ = 0.24). Modest correlations were also observed for overall tapping speed (ρ = 0.20).

**Figure 5.**
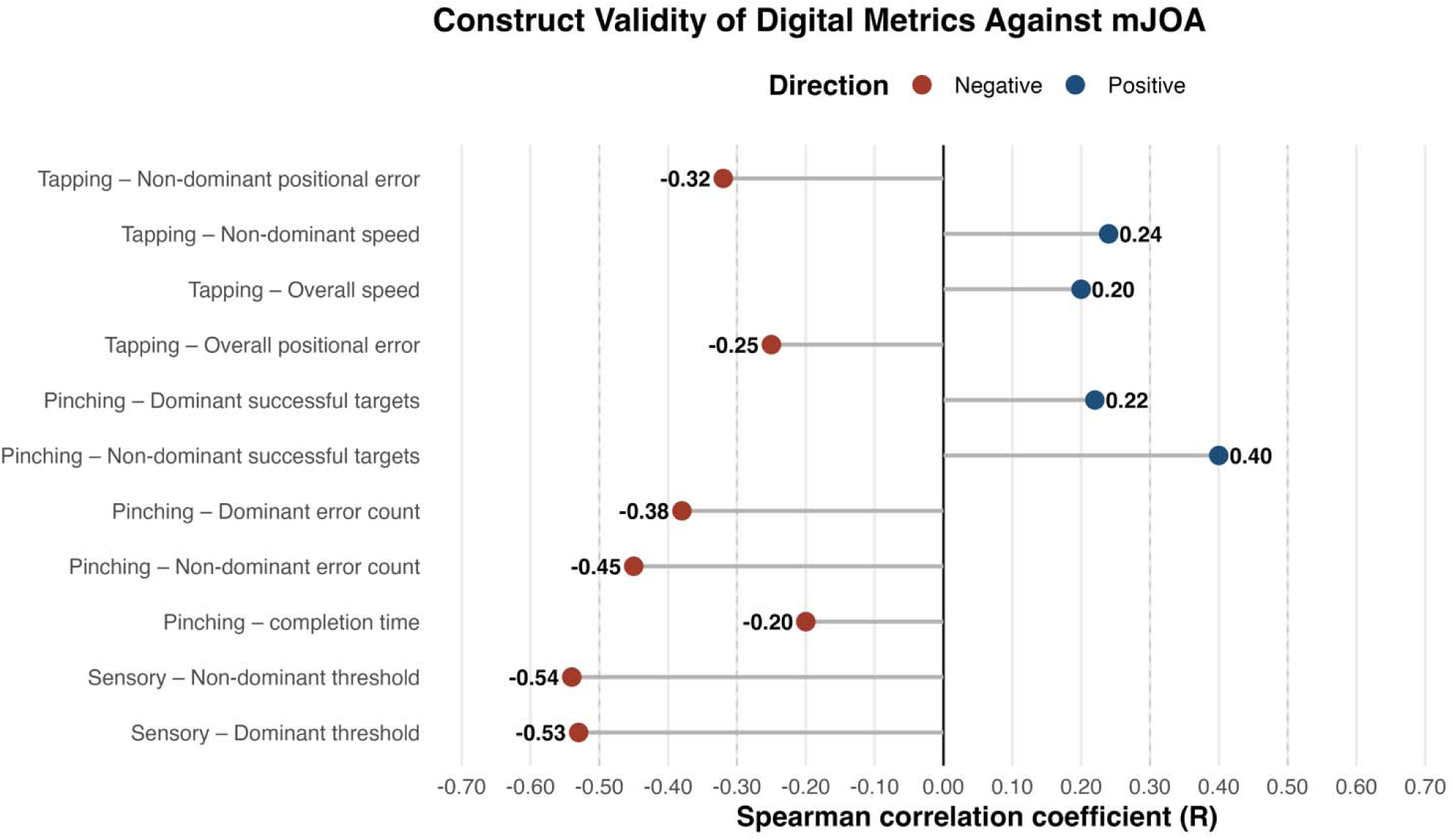
Construct validity of SynapTrack digital biomarkers against the modified Japanese Orthopedic Association (mJOA) score. Spearman correlation coefficients (ρ) of each candidate digital metric with the total mJOA score. Pinching metrics reflect Level 2 outputs. Blue points indicate positive correlations; red points indicate negative correlations. mJOA = modified Japanese Orthopedic Association score.

Pinching-derived digital biomarkers demonstrated evidence of construct validity relative to CSM disease severity (Figure 5). The most informative metrics were those reflecting task success and precision, namely the number of successful targets and error targets. In the more challenging Level 2 non-dominant condition, the number of error targets demonstrated a moderate inverse association with mJOA total score (ρ = −0.45), while the number of successful targets (ρ = 0.40) showed a corresponding positive association, indicating that worse neurological impairment was associated with reduced task success and greater error. These findings suggest that pinching-based digital metrics capture clinically meaningful upper-extremity function relevant to CSM, with higher-complexity task conditions potentially more sensitive to neurological impairment.

The sensory test also demonstrated strong construct validity relative to patient-reported CSM severity (Figure 5). Higher vibratory detection thresholds were consistently associated with worse overall neurological impairment as measured by the mJOA (dominant: ρ = −0.53; non-dominant: ρ = −0.54).

### Known-Groups Validity and Discriminative Performance

Selected SynapTrack digital metrics demonstrated meaningful discrimination between patients with CSM and healthy controls (Figure 6). Among tapping metrics, non-dominant tapping speed showed good discrimination (AUROC = 0.76, 95% CI: 0.68–0.85), with lower speed observed in patients with CSM compared with controls (4.30 ± 1.05 vs 5.28 ± 0.93). Overall tapping speed also distinguished CSM patients from controls (AUROC = 0.71, 95% CI: 0.62–0.80), again reflecting slower tapping performance in the CSM cohort. Pinching metrics also showed evidence of known-groups validity, particularly during the more challenging Level 2 dominant-hand task. The number of successful targets discriminated CSM patients from controls (AUROC = 0.78, 95% CI: 0.62–0.94), with fewer successful targets among patients with CSM (5.86 ± 3.19 vs 8.53 ± 1.08). Completion time likewise demonstrated moderate discrimination (AUROC = 0.69, 95% CI: 0.51–0.88), with patients with CSM requiring longer task completion times compared with controls (46.22 ± 9.56 vs 39.94 ± 5.97 seconds). Finally, the non-dominant vibratory detection threshold demonstrated good discrimination between CSM patients and controls (AUROC = 0.77, 95% CI: 0.64–0.90), with higher thresholds among CSM patients (0.23 ± 0.14 vs 0.14 ± 0.06), consistent with worse sensory detection. AUROC values for all evaluated candidate metrics are shown in Figure 6. Collectively, these findings support the ability of selected SynapTrack metrics to distinguish patients with CSM from healthy controls.

**Figure 6.**
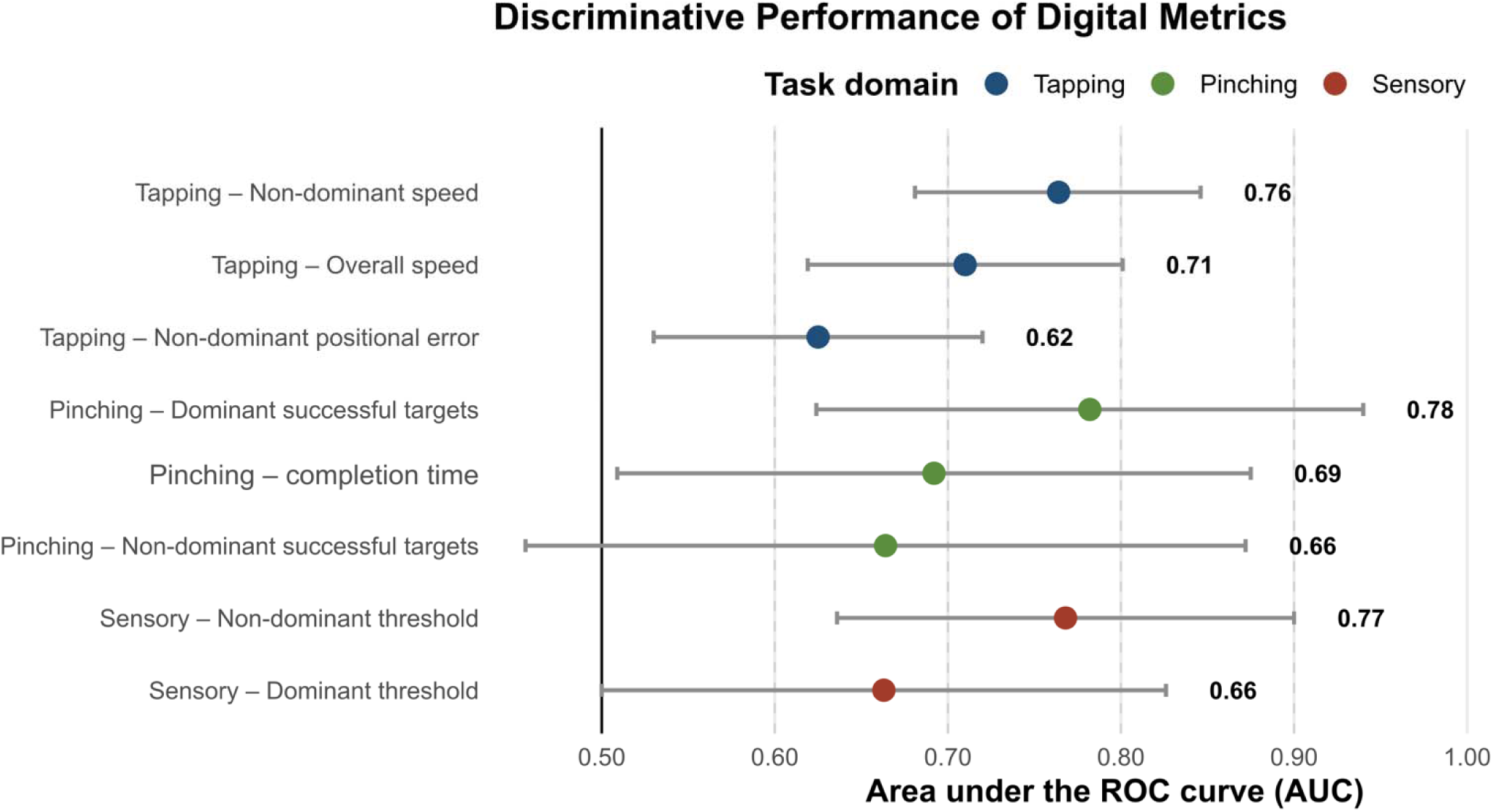
Discriminative performance of SynapTrack digital metrics for distinguishing patients with CSM from healthy controls. Pinching metrics reflect Level 2 outputs. Area under the receiver operating characteristic curve (AUROC) with 95% confidence intervals is shown for each candidate digital biomarker, colored by task domain. The reference line at AUROC = 0.50 corresponds to chance performance.

### Test-Retest Reliability

Selected SynapTrack digital metrics demonstrated good-to-excellent test-retest reliability across sensory, tapping, and pinching domains (Figure 7). Reliability estimates reflect a single-measures ICC comparing an early and a late assessment, with each assessment averaged over two testing days to reduce day-to-day variability. The strongest reliability was observed for the smartphone-based vibratory detection threshold measure, which demonstrated excellent stability across repeated home assessments (ICC = 0.92).

**Figure 7.**
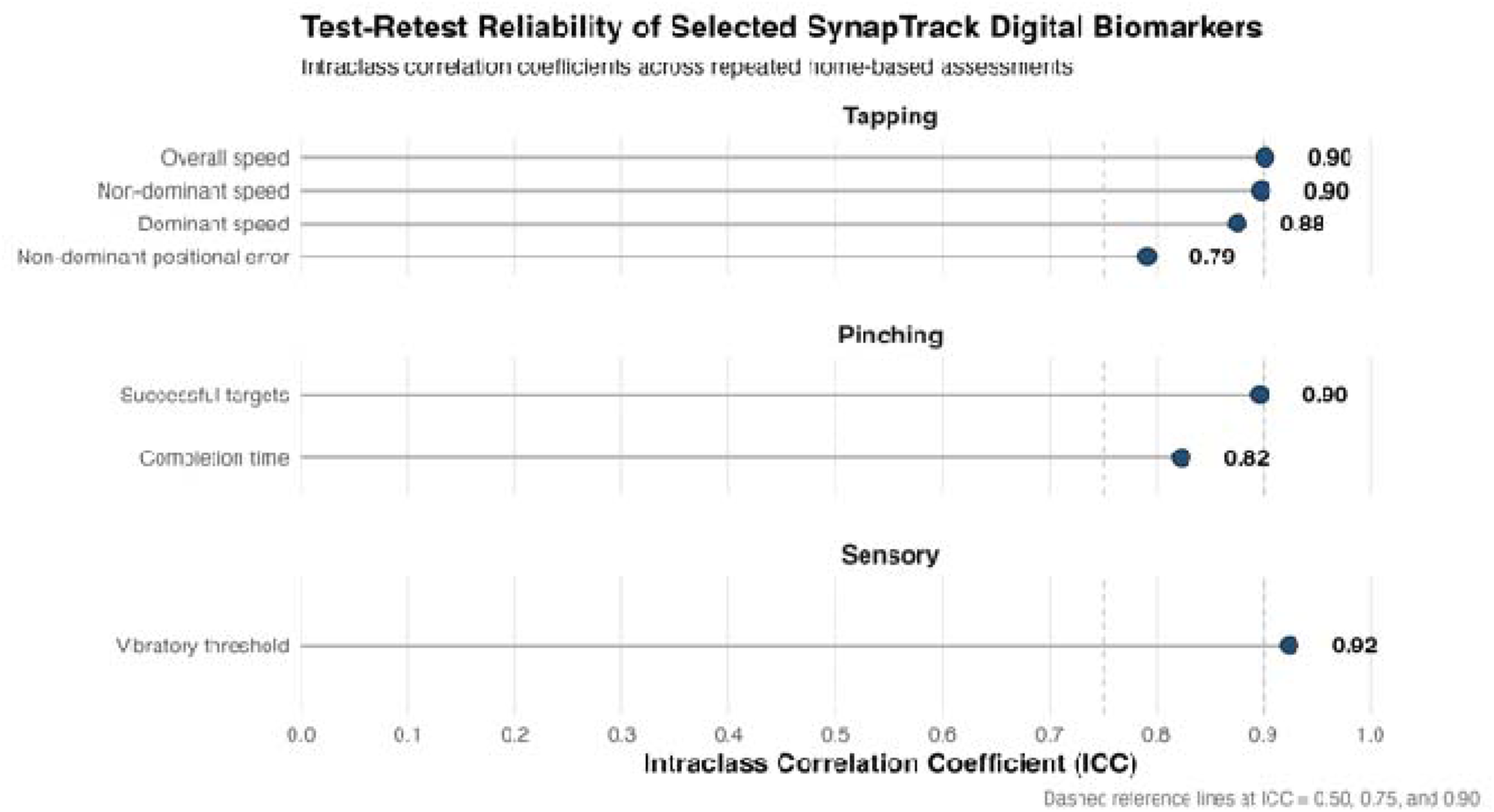
Test-retest reliability of selected SynapTrack digital biomarkers. Intraclass correlation coefficients (ICC [2,1], two-way random-effects absolute-agreement), for tapping, pinching, and vibratory metrics were derived from repeated home-based assessments between an early and a late home-based assessment, with each assessment averaged over two testing days. Dashed reference lines indicate conventional thresholds for moderate (0.50), good (0.75), and excellent (0.90) reliability. ICC = intraclass correlation coefficient.

Tapping-derived biomarkers also showed excellent reliability, particularly speed-based metrics, including overall tapping speed (ICC = 0.90), non-dominant tapping speed (ICC = 0.90), and dominant tapping speed (ICC = 0.88). Non-dominant tapping positional error demonstrated good reliability (ICC = 0.79). Pinching-derived metrics likewise demonstrated generally strong reliability for performance-based measures, including the number of successful targets (ICC = 0.90) and completion time (ICC = 0.82).

## DISCUSSION

In this single-center prospective cohort study, we performed a preliminary evaluation of the reliability and validity of smartphone-derived digital biomarkers of hand sensorimotor function in patients with CSM and healthy controls. To our knowledge this represents the largest cohort digital-biomarker validation study in CSM, providing reasonably precise estimates of validity across multiple sensorimotor domains. Across dexterity tasks (simple tapping and pinching) and a novel haptic vibratory detection task, candidate SynapTrack metrics demonstrated good-to-excellent test–retest reliability, convergent validity against established dexterity measures, construct validity against the mJOA, and ability to discriminate patients with CSM from healthy controls. The most reliable measures – including non-dominant and overall tapping speed, pinching success (number of successful targets), and the vibratory detection threshold – reached ICCs of 0.90 or higher. These values are comparable to or higher than the reliability reported for mature smartphone platforms in multiple sclerosis and Parkinson’s disease.^24,27^ These findings support the feasibility of a multi-domain, smartphone-based platform for objective, remote assessment of upper-extremity neurological function in CSM, and identify a set of candidate digital metrics with sufficient measurement properties to warrant further development.

### Objective measurement of hand function in CSM

The development and testing of digital biomarkers in CSM is motivated by the lack of scalable, validated, and reproducible objective measures to monitor neurological abilities in a disease characterized by spinal cord dysfunction.^34^ In a systematic review of 148 prospective CSM studies, 92% of all outcome measures were questionnaires, and only 8% involved objective physical testing of neurological function such as strength, gait, dexterity, or sensation.^35^ The most widely used scale, the mJOA score, classifies severity in coarse, ordinal gradations, and despite good total-score inter-rater reliability, is prone to misclassification and variability in assessment.^36,37^ Recently, objective tools like the GRASSP-M and GAITRite mat have been developed and have shown quantifiable hand and gait deficits, respectively, tied to disease severity.^19,38^ Such measures show meaningful promise, but implementation barriers, particularly in the context of home testing, have limited broader use. This study’s results indicate that brief, patient-administered smartphone tasks can capture clinically meaningful upper-extremity impairment in CSM with reproducibility approaching that of laboratory reference tests.

This work extends a small but growing body of digital-biomarker research specific to CSM. Rosenthal et al. demonstrated that a tablet-based fine-motor application could distinguish 28 myelopathic patients from age-matched controls with excellent intra-rater reliability and correlations with mJOA reaching r = 0.71 for the strongest task.^39^ More recently, Raju et al. used smartphone video and computer-vision tracking of the grip-and-release test to discriminate CSM from controls and showed that video-derived kinematics correlated with MRI measures of cord compression.^40^ Similarly, Ibara et al. applied machine learning to smartphone videos of the 10-second grip-and-release test, discriminating CSM from controls and demonstrating correlation with JOA-based severity.^41^ Whereas these systems focused on tablet- or camera-based measurements, SynapTrack leverages direct touchscreen interaction together with the smartphone’s built-in haptic and inertial sensors, enabling fully self-administered assessments that may be more practical for unsupervised home use. Critically, no prior CSM-specific platform has integrated objective dexterity and sensory testing in a single self-administered application, and to our knowledge none has applied smartphone haptics to vibratory sensation in this population.

### Digital measurement of dexterity (tapping and pinching)

The strongest convergent validity in our dexterity tasks was observed for tapping speed, which correlated moderately to strongly with the Box and Block Test (ρ = 0.52–0.60) and inversely with 9-Hole Peg Test completion time (ρ = −0.30 to −0.53), and which also showed good discriminative performance (non-dominant tapping speed AUROC = 0.76). Finger tapping is among the most established and widely digitized measures of upper-limb motor function, and our findings are consistent with broader digital-biomarker literature. Specifically, in Parkinson’s disease and multiple sclerosis, tapping features have been shown to correspond with clinician ratings and reliably separate patients from controls.^42–44^ The reliability of speed-based tapping metrics in our cohort (ICC up to 0.90) parallels the good-to-excellent reliabilities reported for tapping features after multi-day aggregation in the Roche PD platform.^27^ The modest correlation between tapping metrics and the mJOA likely reflects a combination of known shortcomings in the patient-reported mJOA and also the reality that a single digital metric may not fully capture all aspects of CSM severity. At the same time, the moderately strong discrimination suggests that tapping features may complement other digital tests to help inform overall CSM assessments, particularly if they show responsiveness to changes in disease severity over time.

The pinching task contributed complementary information. Success-based metrics (number of successful targets) showed consistent moderate convergent validity with the Box and Block Test. Error-based metrics paralleled this pattern, with the number of error targets showing moderate inverse association with mJOA total score (ρ = −0.45) in the Level 2 non-dominant condition. Such results suggest that reduced task success and increased error both index disease severity. The observation that the higher-complexity (Level 2) conditions were more sensitive to disease severity is consistent with the principle that more demanding motor tasks reduce ceiling effects, a pattern previously noted in digital assessments of Parkinson’s disease.^45,46^ The performance-based pinching metrics likewise demonstrated good-to-excellent test–retest reliability (ICC = 0.82 for completion time to 0.90 for the number of successful targets), comparable to the tapping metrics. These results suggest that prioritizing Level 2 task completion may optimize the efficiency of future testing. Separately, we have recently made several technical updates to the pinching task to clarify both task instructions and ease of completion, which we expect will further improve reliability, and likely also its construct and convergent validity.

### A novel smartphone vibratory threshold for an under-measured sensory domain

The vibratory detection threshold was among the strongest-performing biomarkers in this study. It demonstrated excellent test–retest reliability (ICC = 0.92), strong convergent validity against the Box and Block Test (ρ up to −0.65), moderate criterion validity against conventional tuning-fork testing (ρ = −0.44 to −0.46), strong construct validity against mJOA (ρ = −0.53 to −0.54), and good discrimination of patients from controls (AUROC = 0.77). This result is notable because dorsal-column sensory dysfunction is a hallmark of CSM yet remains inadequately captured by conventional clinical assessment. Although altered hand sensation has been reported to demonstrate relatively high sensitivity (76%) and specificity (90%) in case-control studies,^47^ prospective studies have shown poor diagnostic utility of bedside sensory testing across modalities.^48^ Soufi et al. identified no studies that specifically assessed the dorsal-column sensory pathway among 148 prospective CSM cohorts.^35^ Our findings provide preliminary evidence that this clinically important but historically neglected domain can be measured objectively and reproducibly with commercial smartphones.

Our approach builds directly on emerging work using smartphone haptics for sensory testing. Adenekan et al. developed a smartphone-based vibration testing application that uses an adaptive staircase algorithm with multiple reversal points to estimate vibration perception thresholds in patients with pre-diabetes and diabetes.^49^ We adopted the same reversal-based principle for our redesigned vibratory task. Consistent with the findings of Adenekan et al., who reported a moderate correlation between smartphone-derived thresholds and Rydel-Seiffer tuning fork scores (Rs = −0.43),^49^ our redesigned task demonstrated comparable criterion validity while also achieving excellent test–retest reliability (ICC = 0.92). In contrast, an earlier non-adaptive iteration of this task in our pilot testing showed only moderate reliability (ICC ≈ 0.40), underscoring the gain in measurement precision afforded by the staircase design.

### Validity against imperfect reference standards

Several construct-validity correlations against the mJOA were modest to moderate and warrant additional consideration. The mJOA is a coarse ordinal composite spanning multiple neurological domains, whereas our tasks measure only upper-extremity sensorimotor function. This point echoes Lipsmeier et al. in Parkinson’s disease, who attributed weaker sensor–clinical correlations partly to the absence of conceptually comparable clinical items and to limitations of the clinical comparator itself.^27^

The magnitudes we observed are also consistent with those reported for established platforms. In particular, the Roche PD Mobile Application v2 reported sensor–MDS-UPDRS correlations spanning ρ = 0.12–0.71 and characterized these as evidence of preliminary validity.^27^ Notably, those correlations were observed against a “gold standard” clinical reference (MDS-UPDRS) that incorporated objective clinician-administered motor testing. Framed this way, our construct-validity findings are consistent with meaningful convergence against imperfect and only partially overlapping reference standards. Nonetheless, further work is needed to better establish the clinical role of digital biomarkers in CSM, including prospective, longitudinal testing.

### Aggregating repeated assessments improves reliability

Reliability estimates in our cohort were derived from aggregated performance over 2 days of testing, consistent with prior digital-biomarker work showing that sensor-based finger-tapping reliabilities increase when data are aggregated across repeated administrations.^50,51^ This reflects the general measurement principle that aggregation attenuates day-to-day intra-individual variability and captures biological temporal variability.^52^ Thus, in CSM, a remote monitoring schedule capturing multiple short sessions is likely to yield more stable estimates of a patient’s true functional status.

### Limitations

This study has several limitations. First, this was a single-center cohort that was predominantly White, which may limit generalizability to other settings and populations. Second, the test–retest reliability was conducted in a subset of participants, and reliability estimates from modest samples carry wide confidence intervals that we report alongside point estimates. Third, although participants mostly used their own smartphones, device heterogeneity in screen size, sensor characteristics, and haptic-motor specifications is a potential source of measurement variance.^53,54^ Fourth, our analyses were cross-sectional with respect to validity and based on short-interval repeat testing for reliability; we did not assess longitudinal responsiveness, which is a necessary next step.

### Conclusion

In a cohort of patients with CSM and healthy controls, a smartphone-based battery of hand sensorimotor tasks demonstrated good-to-excellent test–retest reliability and preliminary convergent, construct, and known-groups validity. The strongest measures, tapping speed and a novel adaptive-staircase vibratory detection threshold, achieved excellent reliability (ICC ≥ 0.90) and meaningful associations with established dexterity tests and disease severity. Furthermore, the vibratory threshold provides the first objective, reproducible, remotely administered measure of a sensory domain that has been largely overlooked in CSM research. These findings establish the preliminary reliability and validity of SynapTrack and support the feasibility of a multi-domain smartphone platform for objective, remote assessment of upper-extremity neurological function in CSM. Further testing in multicenter cohorts and demonstration of longitudinal responsiveness are now required to determine whether these digital metrics can ultimately improve early detection, monitoring, and surgical decision-making in CSM.

## Data Availability

Due to concerns related to participant privacy, full data are not available, but reasonable requests for data access will be accommodated when possible.

